# Lower probability and shorter duration of infections after Covid-19 vaccine correlate with anti-SARS-CoV-2 circulating IgGs

**DOI:** 10.1101/2021.09.24.21263978

**Authors:** Chiara Ronchini, Sara Gandini, Sebastiano Pasqualato, Luca Mazzarella, Federica Facciotti, Marina Mapelli, IEO Covid Team, Gianmaria Frige’, Rita Passerini, Luca Pase, Silvio Capizzi, Fabrizio Mastrilli, Roberto Orecchia, Gioacchino Natoli, Pier Giuseppe Pelicci

## Abstract

The correlation between immune responses and protection from SARS-CoV-2 infections and its duration remains unclear. We performed a sanitary surveillance at the European Institute of Oncology (IEO) in Milan over a 17 months period. Pre-vaccination, in 1,493 participants, we scored 266 infections (17.8%) and 8 possible reinfections (3%). Post-vaccination, we identified 30 infections in 2,029 vaccinated individuals (1.5%). We report that the probability of infection post-vaccination is i) significantly lower compared to natural infection, ii) associated with a significantly shorter median duration of infection than that of first infection and reinfection, iii) anticorrelated with circulating antibody levels.

## Introduction

SARS-CoV-2 pandemic has resulted in more than 220 million infections and 4.5 million deaths worldwide (Worldometer COVID-19 coronavirus pandemic. https://www.worldometers.info/coronavirus). SARS-CoV-2 vaccination induces strong humoral [1, 2] and cellular [3] immunity and its high effectiveness has been shown in different contexts and populations [4-9]. Knowing the duration and extent of the protection from SARS-CoV-2 infection in individuals who have recovered from COVID-19 or have received the SARS-CoV-2 vaccination is critical to determine the future dynamics of SARS-CoV-2 circulation and have direct impacts on non-pharmaceutical interventions, public health control measures and vaccination strategies. These pieces of information, however, are still an open issue.

### Study design

We performed systematic sanitary surveillance of the personnel working at the European Institute of Oncology (IEO), a large comprehensive cancer center in Milan, Northern Italy. Starting from April 2020, all workers, including health-care, support staff, administrative and research personnel, were tested for SARS-CoV-2 infection by quantitative PCR (qPCR) detection of viral genes, using the Allplex SARS-CoV-2 Assay (Seegene) on nasopharyngeal or saliva samples. Humoral immunity was measured by testing levels of IgGs against the receptor binding domain (RBD) of the spike protein using an in-house ELISA assay [10]. Our assay showed high sensitivity (95.2 %) and specificity (97.6%), that allowed monitoring IgG levels over time in healthy people as well as in Covid-19 patients with accuracy and reproducibility (see Supplementary Materials and Methods for details and [10]). 1,493 participants were initially enrolled into the study starting from April 2020 and monitored before the vaccination campaign, which started on January 7^th^, 2021. 2,029 individuals, including the first cohort, were then vaccinated and monitored until June 2021 (characteristics of our study cohorts are reported in Table S1; timing of tests is described in Supplementary materials – ‘Procedures’ section and Supplementary Figure 1).

### SARS-CoV-2 infections or re-infections prior to vaccination

In the pre-vaccination phase of our screening, we detected 266 SARS-CoV-2 infections (17.8%, 266/1,493). Multivariate logistic models were used to identify independent variables associated with infections during follow-up. While age, COVID-19-symptoms pre-study or a positive PCR-test at baseline did not show significant association, nurse/physician vs. other professionals (researchers, technicians, administratives) were highly correlated with increased probability of infection (P<0.0001). Notably, subjects that were IgG+ at the time of enrollment (T0; Supplementary Figure S2) had 66% significantly lower probability of having a positive swab (OR=0.34, 95%CI: 0.14-0.80, P=0.014, Supplementary Table S4).

We also observed 8 putative re-infections (8/266; ∼3%) (Supplementary Table S2). Re-infections were defined as two PCR-positive samples interspersed with >1 PCR-negative sample. 5 individuals (all IgG+) had reinfection at >60 days. 7 of the 8 re-infected subjects were IgG+ at the time of enrollment (T0; Supplementary Figure S2). Frequency of re-infection with respect to the status of IgG at time of enrollment was ∼9% (7/80) in the IgG+ and 25% (1/4) in the IgG-subjects (difference is not statistically significant, Fisher exact test P=0.335; Table 2). 6 (4/5 IgG+) showed qPCR-positivity to only 1 of the 3 viral-genes tested and with Ct cycles >30. When considering only individuals testing positive for more than one SARS-CoV-2 gene in the PCR assay, frequencies of re-infection decreased significantly (2/266, <1%; 3% vs 0% for IgG+ vs IgG-).

### SARS-CoV-2 infections in vaccinated subjects

2,029 subjects were tested post-vaccination with the Pfizer BNT162b2 or Astra Zeneca (AZ) vaccines. 90% subjects completed the two doses of BNT162b2, and 181 received a single or double dose of AZ. We observed a high rate of vaccination effectiveness, as measured by circulating anti-SARS-CoV-2 RBD IgGs 1 week post-vaccination, with: i) high antibody levels in the entire population (median ∼5 fold increased over the threshold; min=1 and max=12.5) and across each age-group (age range: 19-81y/o); and ii) only 1.9% (39/2,029) of non-responders (IgG levels <0.28) (Figure 1). IgG levels inversely correlated with age, with the lowest levels (median of 7.9) in subjects >70 (median of 20.0 in the age group 19-29; Figure 1). Moreover, levels of IgG monotonically declined over time post-vaccination, though 95.3% (1303/1367) or 98.4% (1030/1047) of tested individuals showed IgG levels above the threshold at 3 or 4 months post-vaccination, respectively (median of 2.22 and 1.57, respectively; Figure 1).

**Figure 1.**
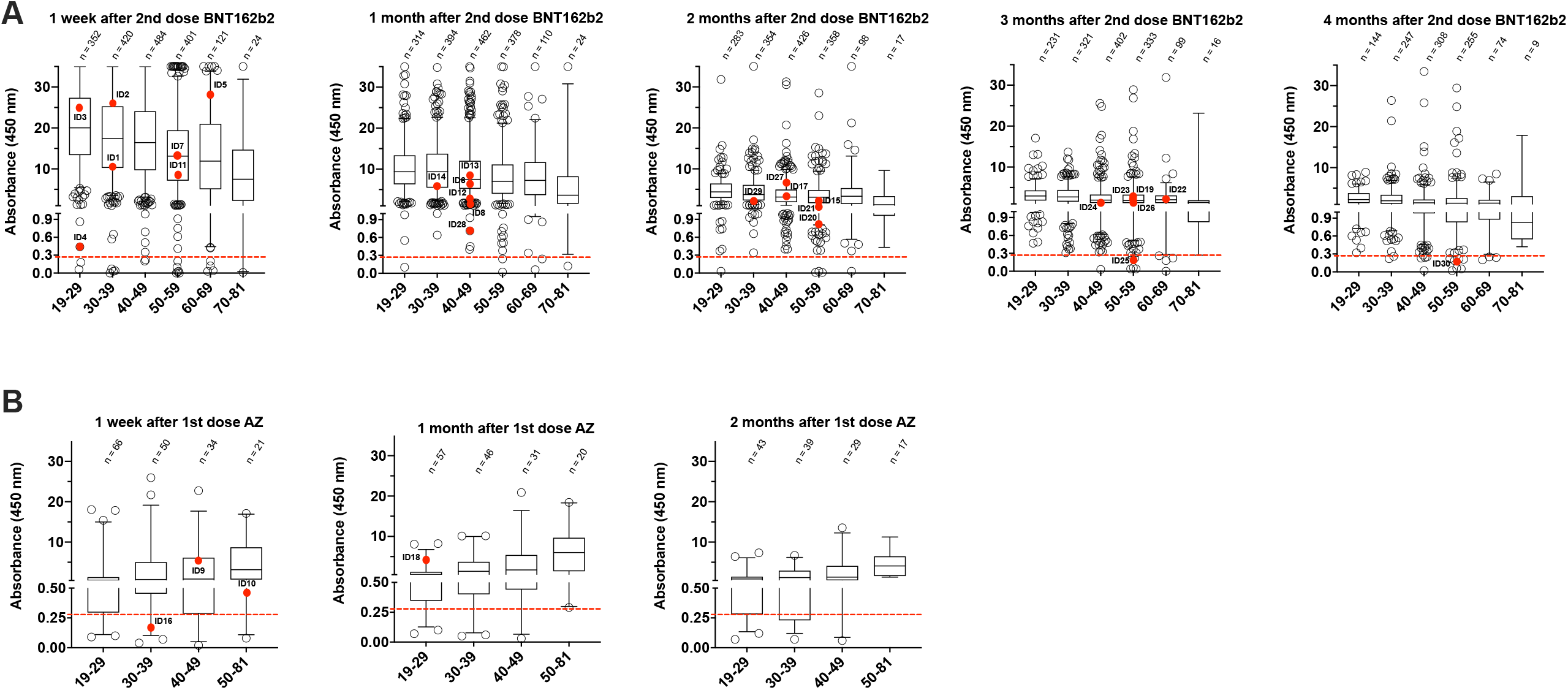
IgG levels against the receptor binding domain (RBD) of the SARS-CoV-2 protein after 2 doses of BNT162b2 vaccine. Individuals are divided by age groups. The red dots highlight the IgG levels in individuals that resulted positive to SARS-CoV-2 infection by qPCR. The dashed red line indicates the threshold of positivity for our serological test (positives>0.28). Boxes define the 25th and the 75th percentiles; horizontal line within the boxes indicates the median and whiskers define the 5th and the 95th percentiles. B, as for panel A after 1 dose of AstraZeneca (AZ) vaccine.

In the 2,029 vaccinated subjects, we identified 30 cases (1.5%) of molecularly-detectable infections (Table 1B). 4 had received only one dose of the AZ vaccine, while all others had completed the two doses of the BNT162b2. Notably, the probability of infection after vaccination was significantly lower than in the non-vaccinated subjects (1.47% vs 9.52%; P<0.0001; Table 2), confirming the effectiveness of vaccination [4-9]. Infections were detected in all age groups except for the oldest (median 47.4 years old; min 23 and max 62; Table 1B). Time of infection varied from few days post-vaccination to >4 months after completion of the vaccination protocol (min 5 days, max 139, median 55 days post-vaccination, Table 1B).

**Table 1A:**
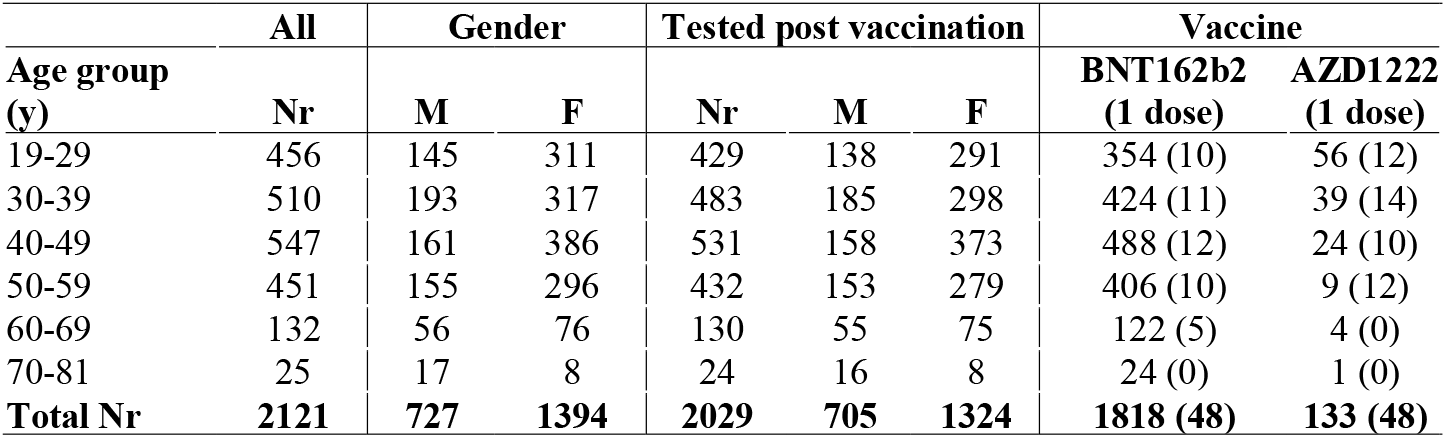
study population.

| Age group (y) | All | Gender |  | Tested post vaccination |  |  | Vaccine |  |
| --- | --- | --- | --- | --- | --- | --- | --- | --- |
|  | Nr | M | F | Nr | M | F | BNT162b2 (1 dose) | AZD1222 (1 dose) |
| 19-29 | 456 | 145 | 311 | 429 | 138 | 291 | 354 (10) | 56 (12) |
| 30-39 | 510 | 193 | 317 | 483 | 185 | 298 | 424 (11) | 39 (14) |
| 40-49 | 547 | 161 | 386 | 531 | 158 | 373 | 488 (12) | 24 (10) |
| 50-59 | 451 | 155 | 296 | 432 | 153 | 279 | 406 (10) | 9 (12) |
| 60-69 | 132 | 56 | 76 | 130 | 55 | 75 | 122 (5) | 4 (0) |
| 70-81 | 25 | 17 | 8 | 24 | 16 | 8 | 24 (0) | 1 (0) |
| **Total Nr** | **2121** | **727** | **1394** | **2029** | **705** | **1324** | **1818 (48)** | **133 (48)** |

**Table 1B:** SARS-Cov2-positive individuals post-vaccination.

| Subjects | Age range | Days post-vaccination | E gene | RdRP gene | N gene | anti-RBD IgG (range 0.28-35) | IgG quartile (min 1, max 4)* | Vaccine |
| --- | --- | --- | --- | --- | --- | --- | --- | --- |
| ID1 | 36-40 | 7 | n | n | 38.96 | 10.65 | 1 | II jabs BNT162b2 |
| ID2 | 31-35 | 5 | n | n | 35.80 | 25.81 | 4 | II jabs BNT162b2 |
| ID3 | 26-30 | 7 | n | n | 38.32 | 24.67 | 3 | II jabs BNT162b2 |
| ID4 | 21-25 | 11 | 38.98 | n | 36.91 | 0.55 | 1 | II jabs BNT162b2 |
| ID5 | 61-65 | 8 | 31.41 | 35.04 | 33.14 | 27.66 | 4 | II jabs BNT162b2 |
| ID6 | 41-45 | 42 | n | 37.67 | n | 6.10 | 2 | II jabs BNT162b2 |
| ID7 | 45-50 | 12 | 30.29 | 33.73 | 31.92 | 12.11 | 2 | II jabs BNT162b2 |
| ID8 | 41-45 | 40 | 25.35 | 27.27 | 27.15 | 3.28 | 1 | II jabs BNT162b2 |
| ID9 | 41-45 | 23 | n | n | 38,9 | 5.84 | 3 | I jab AZ |
| ID10 | 51-55 | 21 | 22.85 | 25.45 | 24.56 | 0.45 | 2 | I jab AZ |
| ID11 | 56-60 | 46 | 21.76 | 23.78 | 22.76 | 9.48 | 2 | II jabs BNT162b2 |
| ID12 | 41-45 | 53 | 21.4 | 22.79 | 19.69 | 4.57 | 1 | II jabs BNT162b2 |
| ID13 | 46-50 | 55 | 36.58 | 38.78 | 34.4 | 8.64 | 3 | II jabs BNT162b2 |
| ID14 | 36-40 | 53 | 28.77 | 31.61 | 29.85 | 6.76 | 2 | II jabs BNT162b2 |
| ID15 | 51-55 | 67 | 29.06 | 31.89 | 28.65 | 1.21 | 1 | II jabs BNT162b2 |
| ID16 | 31-35 | 21 | 37.16 | n | 34.74 | 0.21 | 1 | I jab AZ |
| ID17 | 46-50 | 72 | n | n | 37.12 | 3.09 | 2 | II jabs BNT162b2 |
| ID18 | 26-30 | 55 | n | n | 37.34 | 4.98 | 4 | I jab AZ |
| ID19 | 51-55 | 98 | 21.05 | 23.17 | 20.56 | 3.25 | 3 | II jabs BNT162b2 |
| ID20 | 46-50 | 98 | 30.03 | 32.52 | 28.61 | 0.69 | 1 | II jabs BNT162b2 |
| ID21 | 51-55 | 91 | n | n | 37.01 | 1.59 | 2 | II jabs BNT162b2 |
| ID22 | 56-60 | 88 | n | 38.37 | n | 2.29 | 3 | II jabs BNT162b2 |
| ID23 | 46-50 | 68 | n | 38.83 | n | 2.88 | 1 | II jabs BNT162b2 |
| ID24 | 46-50 | 98 | n | n | 37.04 | 1.69 | 2 | II jabs BNT162b2 |
| ID25 | 51-55 | 99 | n | n | 36.88 | 0.26 | 1 | II jabs BNT162b2 |
| ID26 | 46-50 | 108 | n | n | 36.90 | 3.69 | 1 | II jabs BNT162b2 |
| ID27 | 41-45 | 75 | 35.89 | 36.94 | 35.87 | 6.50 | 4 | II jabs BNT162b2 |
| ID28 | 46-50 | 78 | n | n | 36.07 | 0.71 | 1 | II jabs BNT162b2 |
| ID29 | 31-35 | 116 | n | n | 37.25 | 3.06 | 3 | II jabs BNT162b2 |
| ID30 | 56-60 | 139 | 12.41 | 15.61 | 11.68 | 0.22 | 1 | II jabs BNT162b2 |

\* quartiles normalized to age and time after vaccination; n, not detectable

**Table 2.**
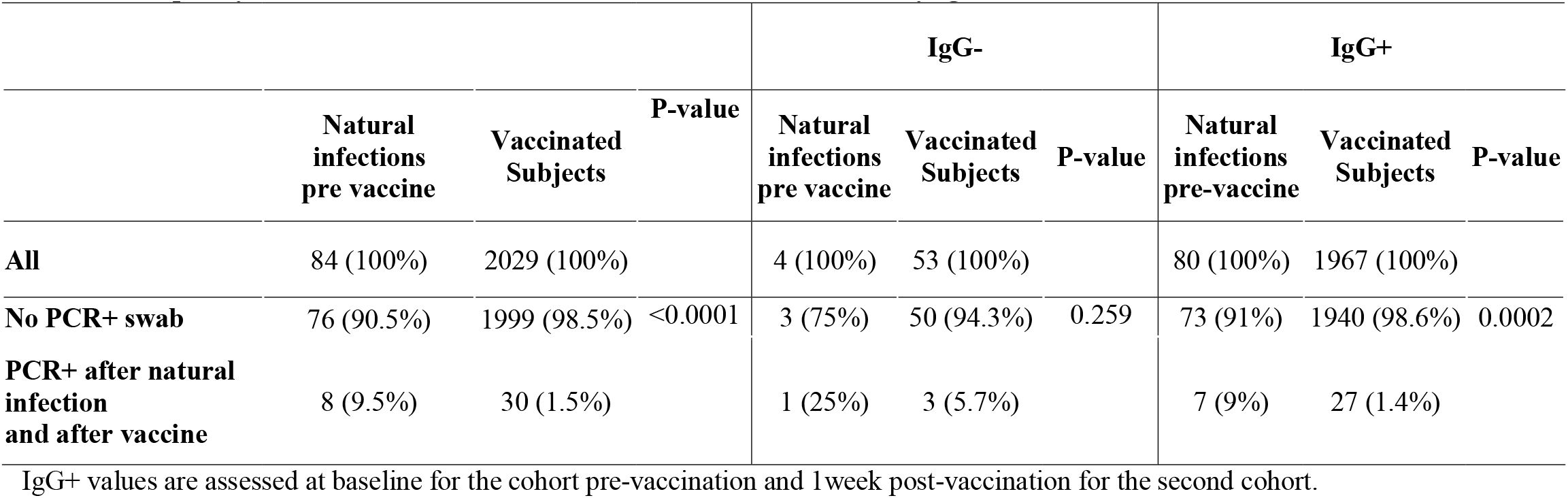
Frequency of natural reinfections and infections after vaccination by IgG status.

The median duration of infections based on a positive PCR test in the vaccinated subjects was 2 days (Interquartile range - IQR: 2-4). Notably, this duration was significantly shorter than post-natural infections (16.5 days; IQR: 11-40.5; P<0.001) or re-infections (11 days; IQR 4-21; P=0.0035) in the pre-vaccinated subjects, suggesting significantly shorter duration of viral shedding in vaccinated individuals as compared to the unvaccinated ones (Table S3 and Figure 2). Moreover, to our knowledge, all infected individuals reported asymptomatic or pauci-symptomatic infections.

**Figure 2.**
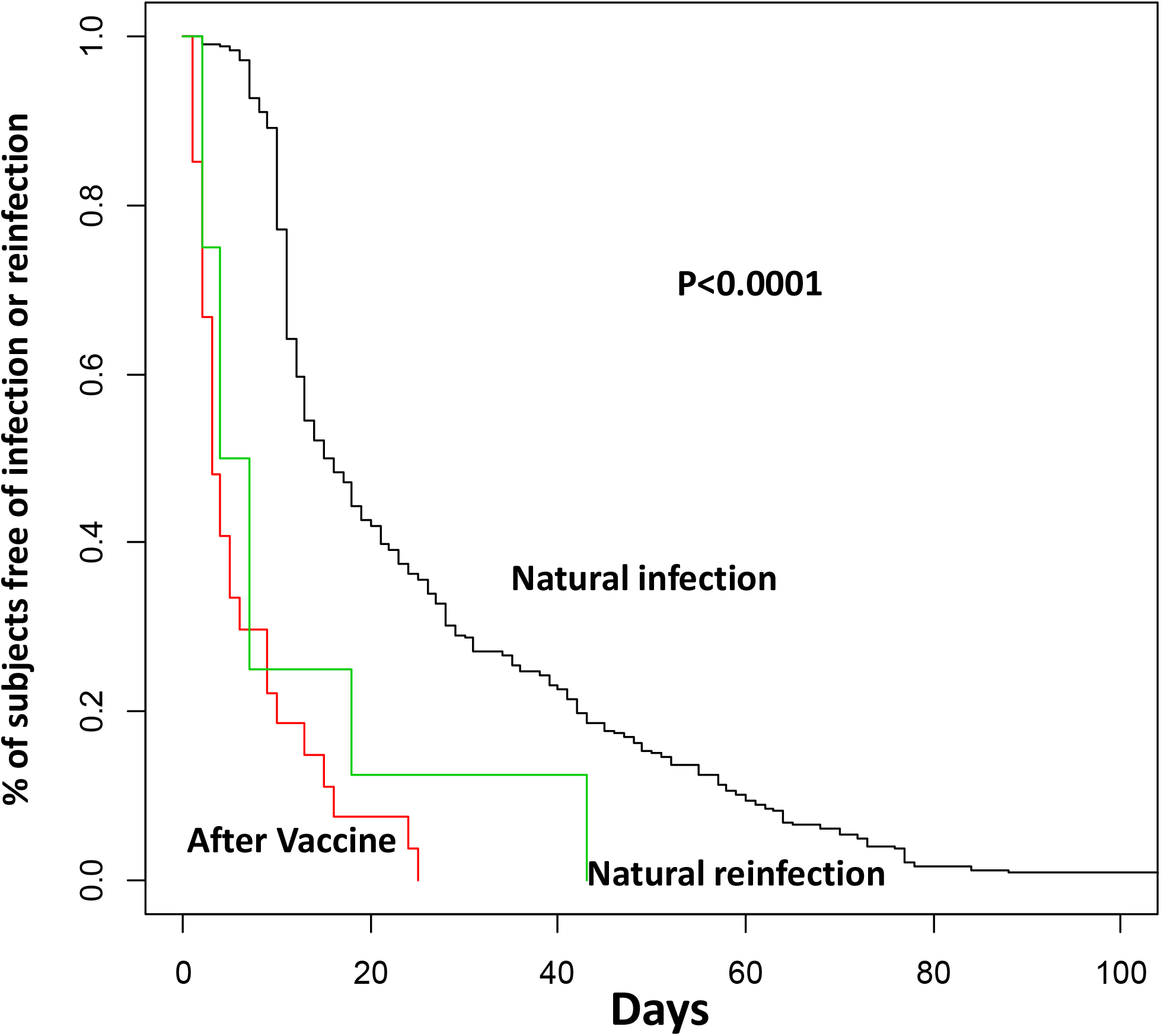
Time of SARS-CoV-2 infections. Kaplan-Meier curves of natural infections (black line); natural reinfections (green line); infections post-vaccine (red line). P-value, Log-rank test.

### Correlations with IgG levels of SARS-CoV-2 infections in vaccine responders

The frequency of molecularly detectable infections among the IgG+ vaccine-responders (subjects that positively responded to vaccination) was significantly lower (1,4%) than in the IgG+ non-vaccinated subjects after natural infection (9%; P=0.0002) and the IgG-vaccine non-responders (5.7%; P=0.042) (Table 2).

Among the newly infected vaccinated-subjects, 3 cases were IgG non-responders (IgG-). Notably, the remaining 27 infected vaccinated individuals were mainly distributed in the lower quartiles of anti-RBD antibody titers (∼74% in quartiles 1 and 2; Table 1B and Figure 1).

## Discussion

Our surveillance study yielded three main findings: i) the probability of infections after COVID-19 vaccine is lower than after natural infection; ii) the few SARS-CoV-2 infections occurring in individuals who mounted a serologically positive response to vaccination are of significantly shorter duration than the first infections in non-vaccinated individuals; iii) the levels of anti-SARS-CoV-2 circulating IgGs were inversely correlated with the frequency and duration of viral detection, as recently reported [11].

This cohort study in healthy workers conducted from the end of the first wave confirmed that reinfection after natural infection is seven times more likely than infection after vaccination. This finding supports the CDC recommendation that all eligible persons be offered COVID-19 vaccination, regardless of previous SARS-CoV-2 infection status. However, the probability of reinfection largely depends on pre-existent IgG positivity. Thus, serological testing in vaccinated individuals might help to identify the population at higher risk of infection.

Reinfections have been reported internationally since June 2020, although they remain uncommon: test results of 4 million people in Denmark found that < 1% of persons who tested positive for SARS-CoV-2 experienced reinfection [12]. The vastly shorter duration of post-vaccine infections likely has major impacts on models to predict epidemiological dynamics, which critically rely on this parameter [13, 14], and may suggest a modification of the isolation policies, which still recommend releasing from isolation 10 days after a first positive PCR test for asymptomatic testing, without distinction for vaccinated subjects [15].

Large longitudinal cohort studies with regular testing are needed to provide systematic epidemiological, virological, immunological, and clinical data useful to understand the rates of reinfection and their implications for health policies. Moreover, data presented in this study will need to be updated to estimate the impact of the delta variant on reinfection/post-vaccine infection risk, considering that the delta variant was not diffuse in our country at the time of testing described in our study.

## Conclusions

Overall, our data show that individuals who responded to vaccination based on the detection of anti-RBD antibodies were still susceptible to SARS-CoV-2 productive infection, suggesting caution, especially for healthcare workers that are daily in contact with fragile patients, such as cancer patients in our Institute. However, the probability of infection after vaccination is rare and significantly less frequent compared to reinfection after natural infection, in particular in responders, which are the vast majority. Furthermore, the duration of infection in vaccinated individuals is significantly shorter to the one observed post-natural infection, suggesting that post-vaccination viral shedding is likely very limited, recommending for a revision of the isolation policies, that could drastically reduce the time of quarantine, with clear important social and economical implications.

## Data Availability

The datasets generated during and/or analysed during the current study are available from the corresponding author on reasonable request.

## Acknowledgements

We thank Fondazione Europea Guido Venosta, National Instruments Corporation, Ralph Lauren and Fondo MFS for funding this research and Fondazione IEO-CCM for fund-raising.

