## Supplementary Material for "Lower probability and shorter duration of infections after Covid-19 vaccine correlate with anti-SARS-CoV-2 circulating IgGs"

**Materials and Methods**

***Study design and participants***

SOS-COV2 is a prospective cohort study including staff working at the European Institute of Oncology in Milan, Italy. All health-care workers, support staff, and administrative staff working at hospital sites, who could provide written informed consent to participate in the study, and anticipated remaining engaged in follow-up for 12 months were eligible. Participants were excluded from this analysis if they did not participate to the screening after enrolment. Recruitment began in May 2020. Ethical approval was granted by the Institutional review board.

***Statistical methods***

We investigated the rate of infection/reinfection by positive status at baseline in the four groups identified by PCR and IgG (PCR- IgG-; PCR+ IgG+; PCR+ IgG-; PCR-IgG+). We collected information also on the values of Ct of genes for positive PCR and we did a further analysis including only reinfections with at least two positive genes. Participants reporting cough, fever, anosmia, or dysgeusia were defined as having COVID-19 symptoms.

We did a multivariable logistic regression analysis to estimate Odds Ratios (ORs) to measure the association between the exposure (positive status at baseline) and infection/reinfection adjusting for significant confounders.

We also carried out a multivariable mixed effects model for repeated measures analyses to investigate factors associated with changes in immunological values. We investigated whether the information at baseline, such as symptoms status at baseline, profession and age, were independently associated with serological values.

***Procedures***

At baseline questionnaires on risk factors of exposures were sent electronically. SARS-CoV-2 antibody testing and real-time PCR (rtPCR) was done at enrolment and at the end of the study. Furthermore, antibody testing was done every 4 weeks. PCR test was done after a positive serological test, in case of symptom, after holidays and every 2 weeks for medical doctors. Swabs were taken by a trained professional (including anterior nasal swabs or combined nose and oropharyngeal swabs). SARS-CoV-2 serology was done with locally validated assays. Testing was done in the clinical laboratory with locally validated testing platforms. COVID-19 vaccination was introduced into this cohort in January, 2021.

Participants were assigned to the positive cohort if they met one of the following criteria: antibody positive on enrolment or a positive PCR result at enrolment. Participants were assigned to the negative cohort if they had a negative antibody test and no documented previous positive PCR or antibody test.

A possible reinfection was defined as a participant with two positive PCR samples with a negative PCR between the two positive PCR samples and considering a positive PCR after 60 or more days, based on previous study. For this second analysis participants with recurrent positive PCR results less than 60 days apart were not considered possible reinfections. We also carried out a supportive viral genomic data from samples available.

Data were collected on potential confounders, including profession and participant demographics, to permit adjustment in analysis.

The cohort susceptible to primary infection (PCR- IgG-): from first antibody-negative date to first positive PCR date or seroconversion (if no positive PCR test had been reported before seroconversion); or if neither of these occurred, to censor date. The cohort with previous infection (PCR+ IgG+; PCR+ IgG-; PCR-IgG+): the earliest date for previous infection was taken as whichever was first of the positive PCR result or the first positive antibody test (IgG>0.28).

The primary outcome was a reinfection in the positive cohort or a primary infection in the negative cohort, determined by PCR tests.

***SARS-COV-2 detection in respiratory specimens***

Nasopharyngeal specimens were collected by trained healthcare professionals, while saliva samples were self-collected by the participants to the study, allowing at least one hour from eating, drinking and/or brushing of teeth before sample collection. Samples were stored at 4°C until use for processing, usually not more than 2 days after collection. Saliva samples were diluted 1:1 with Sputasol (per 100 ml: 0.1 g DTT, 0.78 g NaCl g, 0.02 g KCl, 0.112 g Na_2_HPO_4_, 0.02 g KH_2_PO_4_) and incubated for 5 min at room temperature, shaking at 500 rpm, in order to lose viscosity. For viral RNA extraction both Sputasol-treated saliva samples and nasopharyngeal swabs were inactivated with DNA/RNA shield (Zymo Research, Euroclone). Viral RNA was extracted from 300 ul of inactivated samples using the Sera-Xtracta Virus/Pathogen kit (Cytiva), following the manufacturer’s instructions. Detection of the Sars-CoV-2 viral genes was performed by rt-qPCR using Allplex 2019-nCoV Assay and, more recently, the Allplex Sars-CoV-2 Assay from Seegene, following the manufacturer’s specifications. Amplification of viral gene and data analysis was performed using the CFX96 Rea-time PCR Detection System (Biorad) and the Seegene Viewer platform, respectively.

***Serological tests for SARS-COV-2***

Serological assays for SARS-CoV-2 were conducted as described (Bruni M. et al, J. Clin. Med. 2020, 9(10), 3188). Various commercial assays that utilize distinct viral antigens and detect different antibody classes are available. However, SARS-CoV-2 serological tests available on the market do not always allow systematic simultaneous detection of a wide antibody spectrum for several antigens in a reliable and flexible manner. Conversely, serological enzyme-linked immunosorbent assays (ELISA) to detect immunoglobulins raised against the highly immunogenic receptor binding domain of the viral Spike Soluble Ectodomain (Spike) (RBD) provided robust results in terms of accuracy and reproducibility, that allow monitoring of IgG levels over time in healthy people pre and post-vaccination, as well as in covid-19 patients. Briefly, the recombinant Spike SARS-CoV-2 glycoprotein RBD was produced in mammalian HEK293T cells, purified by affinity chromatography, quantified and stored in liquid nitrogen. To detect immunoglobulins G (IgG) against the SARS-CoV-2 Spike RBD glycoprotein, purified RBD was adsorbed to a Nunc Maxisorp ELISA plate, aspecific binding was blocked by incubation with PBS-BSA 3% before applying patients’ sera to be analyzed. Anti-RBD IgG presence was revealed with secondary anti-human-IgG antibody (BD, clone G18-145) conjugated to HRP by Glomax reading at 450 nm. The assay has been validated with a cohort of n = 56 COVID-19 subjects (severe, moderate and mild disease) and n = 463 (subjects collected in pre-COVID era, between 2012 and 2015). ROC curves have been implemented to determine the sensitivity and specificity of the assay, based on which IgG positivity was defined as absorbance at 450 nm >0.28 with a sensitivity of 95.2 % and a specificity of 97.6% (Bruni M. et al, J. Clin. Med. 2020, 9(10), 3188). To work in the linearity range of the ELISA response, sera after vaccination were diluted either 1:200, 1:900 or 1:3645, and for the sake of clarity the OD at 450 nm was scaled to the 1:200 dilution before plotting.

**Supplementary Figures**

**Figure S1. Workflow of our study pre- and post-vaccination**

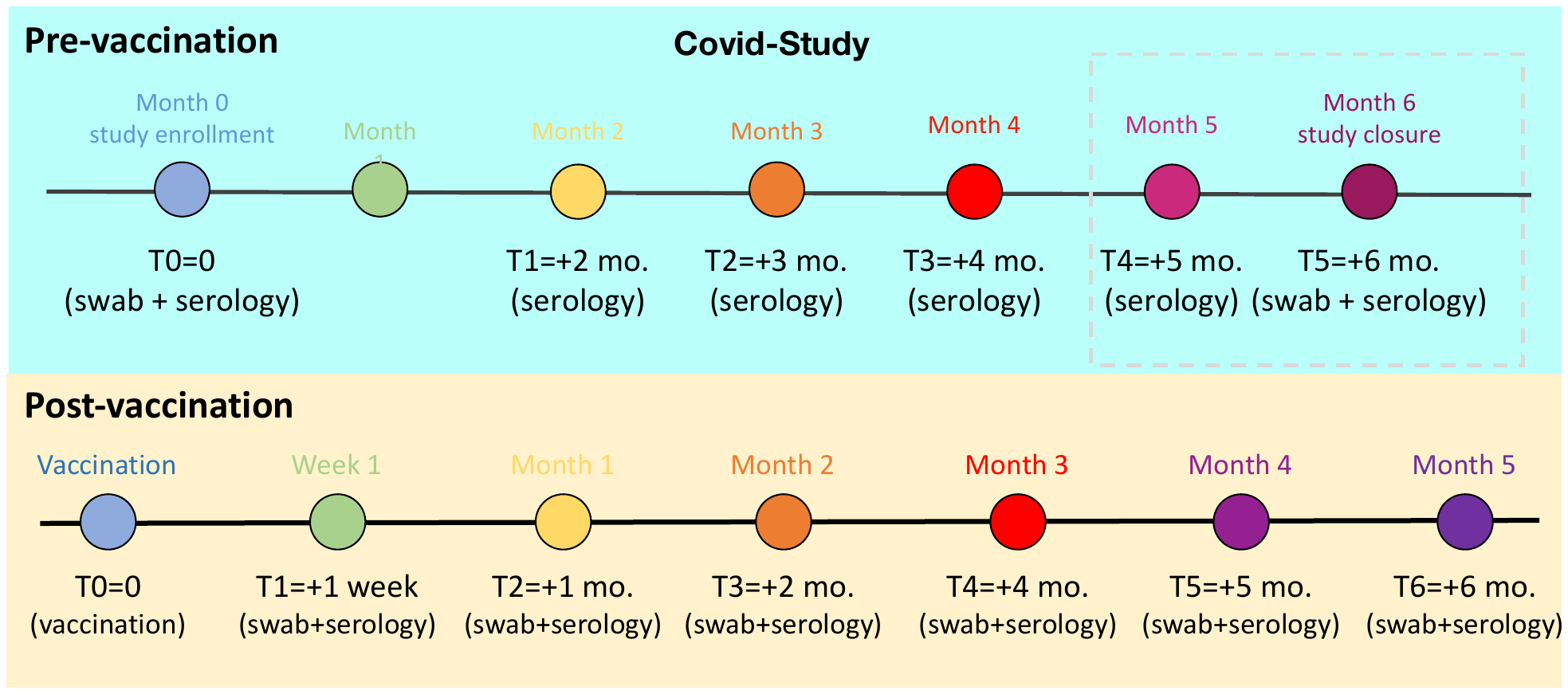

**Figure S2. Study design**

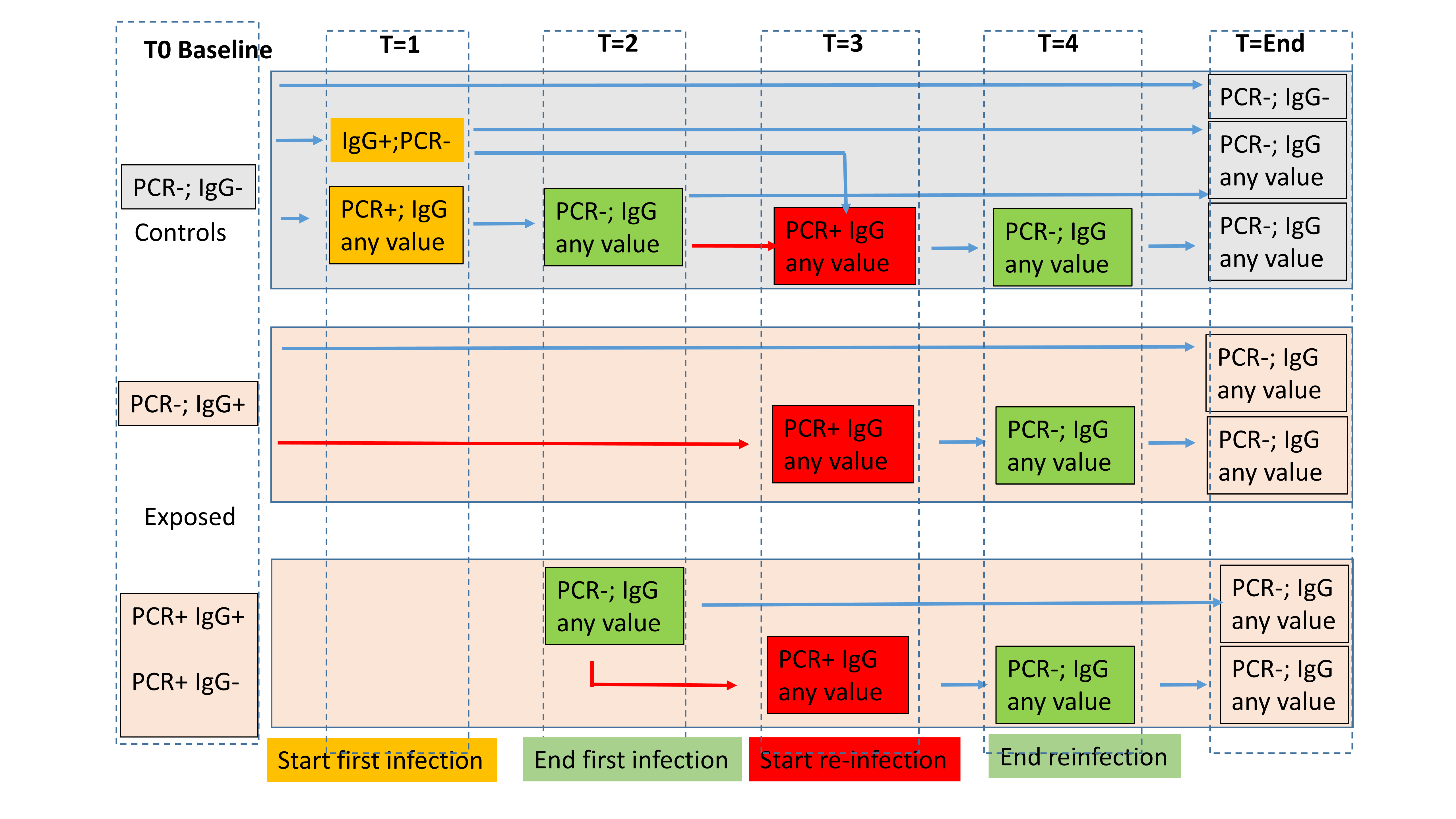

**Supplementary Tables**

**Table S1 Descriptive characteristics of subjects included in pre- and post-vaccination cohorts**

|  |  | **Pre vaccination** | | **Post vaccination** | |
| --- | --- | --- | --- | --- | --- |
|  |  | **Nr**  **(Tot=1493)** | **%** | **Nr**  **(Tot= 2029)** | **%** |
| **Age, Median (Q1-Q3)** |  | 41 (31-49) |  | 42 (31-51) |  |
| **Gender** | Female | 994 | 66.58 | 1324 | 65.25 |
|  | Male | 499 | 34.42 | 705 | 34.75 |
| **Profession** | Administration | 371 | 18.28 | 225 | 15.07 |
|  | Nurses | 310 | 15.28 | 89 | 5.96 |
|  | Researcher | 260 | 12.81 | 250 | 16.74 |
|  | Physician/Clinician | 784 | 38.64 | 703 | 47.09 |
|  | Student | 110 | 5.42 | 63 | 4.22 |
|  | Technician | 165 | 8.13 | 142 | 9.51 |
|  | Other | 29 | 1.43 | 21 | 1.41 |

**Table S2. Features of reinfections (positive swab)**

|  |  |  |  |  |  | **Viral genes at T0** | | | **Viral genes at first infection** | | |
| --- | --- | --- | --- | --- | --- | --- | --- | --- | --- | --- | --- |
| **ID** | **PCR status** | **IgG value** | **Duration first infection** | **Time to second infection** | **Duration second infection** | **E** | **RdRP** | **N** | **E** | **RdRP** | **N** |
| 1 | Negative | 0.45 | 77 | 43 | 7 |  |  |  | 36.78 | n | 37.57 |
| 2 | Negative | 0.68 | 52 | 188 | 4 |  |  |  | n | n | 37.97 |
| 3 | Negative | 0.86 | 41 | 80 | 18 |  |  |  | n | n | 36.49 |
| 4 | Negative | 0.91 | 70 | 220 | 2 |  |  |  | n | n | 37.65 |
| 5 | Negative | 1.18 | 73 | 161 | 2 |  |  |  | 32.3 | 36.68 | 31.25 |
| 6 | Positive | 0.39 | 7 | 34 | 43 | n | n | 38.04 | n | 37.67 | n |
| 7 | Positive | 0.05 | 154 | 50 | 7 | n | 35.99 | 34.68 | n | 37.46 | n |
| 8 | Positive | 0.50 | 108 | 126 | 4 | 31.14 | 32.62 | 31.14 | n | n | 38.8 |

n, not detectable

**Table S3. Duration of infections and time from first and second infection (negativity)**

|  | **Median** | **Lower Quartile** | **Upper Quartile** | **P-value** |
| --- | --- | --- | --- | --- |
| **First infection** | 16.5 | 11 | 40.5 | <.0001 |
| **Reinfection** | 11 | 4 | 21 | 0.0035 |
| **Duration of negativity** | 34 | 21 | 85 |  |
| **Infection after vaccine** | 2 | 2 | 4 | Reference |

**Table S4. Odd Ratio and 95% Confidence Intervals for the association with infection**

|  |  | **OR** | **Low 95%CI** | **Up 95%CI** | **P-values** |
| --- | --- | --- | --- | --- | --- |
| **Age** |  | 1.01 | 1.001 | 1.03 | 0.038 |
| **Symptoms pre study** | **Yes vs No** | 1.37 | 0.96 | 1.96 | 0.087 |
| **Profession** | **Nurse/Phisician vs other** | 1.90 | 1.44 | 2.51 | <.0001 |
| **Swab at baseline** | **Neg vs Pos** | 0.46 | 0.11 | 1.89 | 0.279 |
| **IgG at baseline** | **>28 vs <28** | 0.34 | 0.15 | 0.80 | 0.014 |

OR from multivariable logistic models including information about personal history.
Legend OR=Odd ratio; considering IgG as continuous variable P=0.06. Adjusted for time from baseline.

**Table S5. Descriptive characteristic of subjects included in the study pre-vaccination (n=1493)**

|  | **Categories** | **Nr** | **%** |
| --- | --- | --- | --- |
| **Personal History of Covid-19 Symptoms** | No | 1263 | 84.59 |
|  | Yes | 229 | 15.34 |
|  | Missing | 1 | 0.07 |
| **Flu between February and April 2020** | No | 1057 | 70.8 |
|  | Yes | 201 | 13.46 |
|  | Missing | 241 | 16.14 |
| **Contact with Covid-19 positive subjects in family** | No | 1170 | 78.37 |
|  | Yes | 54 | 3.62 |
|  | Missing | 267 | 17.01 |
| **Contact with Covid-19 positive at work** | No | 677 | 45.34 |
|  | Yes | 549 | 36.77 |
|  | Missing | 267 | 17.88 |
| **Contact with Covid-19 positive extra-work** | No | 1178 | 78.9 |
|  | Yes | 52 | 3.48 |
|  | Missing | 269 | 18.02 |
